# Persistent Symptoms Among Frontline Health Workers Post-acute COVID-19 Infection

**DOI:** 10.1101/2021.12.03.21267225

**Authors:** CN Wose Kinge, S Hanekom, A-L Smith, F Akpan, E Mothibi, T Maotoe, F Lebatie, P Majuba, I Sanne, C Chasela

**Affiliations:** Right to Care, On the Lake, 1006 Lenchen North Avenue Centurion, 0046, South Africa; Stellenbosch University, Faculty of Medicine and Health Sciences, Physiotherapy Division, Fancie van Zijl Drive, Tygerberg, 8000, South Africa; University of the Witwatersrand, Faculty of Health Sciences, School of Clinical Medicine, Clinical HIV Research Unit, South Africa; University of the Witwatersrand, Faculty of Health Sciences, School of Public Health, Department of Epidemiology and Biostatistics, South Africa

**Keywords:** COVID-19 symptoms, Persistent, Long, Frontline health workers, Right to Care, South Africa

## Abstract

**Background:** Growing evidence shows that a significant number of patients with COVID-19 experience prolonged/persistent symptoms, also known as Long COVID. Reports of Long COVID are rising but little is known about prevalence in non-hospitalized patients.

**Objective:** We sought to identify the persistent symptoms of COVID-19 in frontline workers at Right to Care (RTC) South Africa who have past the acute phase of illness with a view to establishing rehabilitation programs for its employees and the community at large.

**Methods:** This was a cross-sectional survey. We analysed data from 207 eligible COVID-19 positive RTC frontline workers who participated in a post-COVID online self-administered survey. The survey was active for two months. Frequencies and median were calculated for categorical and continuous variables, respectively.

**Results:** The survey response rate was 30% (62 out of 207); of the 62 respondents with a median age of 33.5 years (IQR= 30–44 years), 47 (76%) were females. The majority (n=55; 88.7%) self-isolated and 7 (11.3%) were admitted to hospital at time of diagnosis. The most common comorbid condition reported was hypertension particularly among workers aged 45–55 years. Headache, body ache, fatigue, loss of smell, dry cough, fever, and loss of appetite were the most common reported symptoms at time of diagnosis. Persistent symptoms were characterized by fatigue, anxiety, difficulty sleeping, chest pain, muscle pain and brain fog, being the six most reported.

**Conclusion:** The impact of persistent/Long COVID-19 on the health of frontline workers could have direct impact in health service delivery. Given the rise in cases of COVID-19 in South Africa and the world at large, the prevalence of Long COVID is likely to be substantial and therefore need for rehabilitation programs targeted at each of the persistent (Long) COVID symptoms is critical.

## Introduction

Since the outbreak of Coronavirus Disease 2019 (COVID-19) and the unprecedented and rapid spread of the disease, resulting in high morbidity and mortality, ^[1]^ scientific evidence suggests that 10% of patients with COVID-19 experience symptoms beyond 3–4□weeks^[2]^ especially in hospitalized patients.^[3]^ In a study by Huang et al.^[4]^, more than 75% of patients that were previously hospitalized with COVID-19 had at least one lingering symptom six months later. Some suggested residual effects of SARS-CoV-2 infection reported so far include but not limited to fatigue, shortness of breath, joint pain, chest pain, cognitive disturbances, and drop in the quality of life.^[3,4]^ Growing evidence shows that a significant number of patients with COVID-19 experience prolonged/persistent symptoms, also known as Long COVID. Reports of Long COVID are rising but little is known about prevalence in non-hospitalized patients.

South Africa reported its first case of COVID-19 on March 5, 2020,^[5]^ since then, there has been a rise in infection especially among frontline health workers. The novel coronavirus SARS-CoV-2 poses an even greater risk to frontline health workers than any other infectious previously reported. By May 6, 2020, South Africa reported 511 cases in health care workers (HCWs) (7% of the national total), with nurses accounting for 53% of total HCW cases.^[6]^ With each wave of infection, HCWs have continued to experience a high number of infections as well as mortality; and for those that have past the acute phase of infection, they were required to return to work at the minimum time possible.

Health care workers at Right to Care South Africa, a Non-profit Organization operating across seven provinces providing health services, have not been spared with the advent of COVID-19. In this study, we sought to identify the persistent/Long COVID symptoms that frontline workers at Right to Care South Africa, experience post-acute phase of illness.

## Methods

### Design

An anonymous cross-sectional survey was conducted on an online platform from 15 February to 15 April 2021 in South Africa. The survey was reviewed and approved by Right to Care COVID-19 Business Continuity Committee and the Health Research Ethics Committee (HREC) of Stellenbosch University (SU), South Africa (HEA-2020-19255).

### Participants

The study was conducted among Right to Care (RTC) South Africa staff. RTC is a Non-governmental Organization (NGO) working in response to public healthcare emergency including HIV and AIDS, tuberculosis (TB) as well as emerging outbreaks such Ebola and COVID-19. All staff working either on a permanent or short-term basis, across seven provinces (Gauteng, Free State, Eastern Cape, Northern, North-West and Mpumalanga) in South Africa where RTC operates and had tested positive for COVID-19 were eligible to participate in the survey. Participation was voluntary. Staff who had inconclusive, negative, pending, and missing results, as well those with invalid, missing phone and/or emails were excluded. COVID-19 testing for the organization is done at private AMPATH and Lancet laboratories, as well as at the National Health Laboratory Services (NHLS), a government institution. The test results are sent to the designated RTC Medical Officer and captured on the RTC COVID-19 database.

### Survey Instrument

The survey questionnaire was developed by the Right to Care (RTC) Implementation Science (IS) team in partnership with the COVID-19 Business Continuity Committee, and Stellenbosch University research team. It consisted of 67 items covering the following seven aspects: (1) demographics (age, sex, and ethnicity), (2) general health status (smoking, alcohol, and recreational drug use) (3) comorbidities, (4) health status at time of COVID-19 diagnosis, (5) health status post COVID-19, (6) current health status post COVID-19, and (7) physical activity. Participants were asked to rate how bothersome the post COVID-19 symptoms were, using a 10-point end labelled adopted response scale was (least bothersome to most bothersome). Similarly, for the severity of current post COVID-19 symptoms, a 10-point end labelled slider scale was adopted for the response scale (not noticeable to unbearable). An eight-point Likert scale was adopted for the response scale about number of days spent on exercise prior to COVID-19 diagnosis (None to Seven), whereas a nine-point Likert scale was adopted for the response scale on the time (in minutes) spent on exercise (10 to greater than 150) in line with WHO recommended activity levels of 150 minutes moderate intensity exercise per week.^[7]^.

### Data Collection

The survey was captured on REDCap^[8]^ platform with an information page at the beginning to provide for respondents’ consent before they completed the survey. The online self-administered questionnaire was promoted by distributing the survey link through email. The survey link was also circulated to line managers to share with their respective employees that report directly to them who were known COVID-19 survivors. It was also promoted through the Group RTCSA email platform. For those without email addresses, the survey was sent out via SMS and WhatsApp messages by the RTC eHealth Systems and Innovation team. These were done to increase response rate. A total of 129 SMS/WhatsApp messages with survey link were sent out to employees through RTC eHealth Solutions and Innovation team and 73 were sent out via email directly through REDCap. A weekly reminder was sent out via email as well as SMS and WhatsApp for the duration of the survey period. This was active for a two-month period. All participation was voluntary, and responses were anonymous. The collected data were password-protected and only the SU/RTC Implementation Science (IS) researchers had the access code to login for using the data for analysis.

### Statistical analysis

Data management and analysis were conducted using Stata/SE Version 15.1. Descriptive information including the characteristics of respondents, underlying medical conditions, post COVID-19 as well as persistent (Long) post COVID-19 symptoms and physical activity levels were calculated. Long COVID, was defined as the presence of symptoms for four weeks and beyond.^[5]^ Differences between categorical variables were determined using Chi-square or Fisher’s exact tests. A t-test was used to compare the statistical difference in means of days and time spent on physical activity before COVID-19 diagnosis and after recovery. Statistical significance was considered when p-values were <0.05.

## Results

### Demographics

A total of 916 employees were tested for COVID-19 between January 5 2020 and February 7 2021 and captured on the RTC COVID database. Of these, 248 (27%) tested positive, 207 were eligible to take part in the survey, 99 (47.8%) participated, and only 62 (giving a response rate of 30%) consented and completed the survey as shown (Fig. 1). Of the 248 staff who tested positive for COVID-19, approximately 38% (n=95) and 17% (n=42) were community/field health workers and medical/clinical personnel, respectively as shown (Supplementary Table 1). COVID-19 person under investigation (PUI)/suspect was the main reason for testing, accounting for 61% (n=151) of positive cases notably in the Free State (33%; n=81), Mpumalanga (25%; n=61) and Western Cape (28%; n=70) provinces. The highly hit district municipalities were Thabo Mofutsanyane (29%; n=73), Ehlanzeni (23%; n=58), and Overberg (10%; n=25). Staff who tested positive were significantly different (p<0.05) from others (negative, pending, inclusive and missing test results) across cadres, province, districts and reasons for testing.

**Figure 1:**
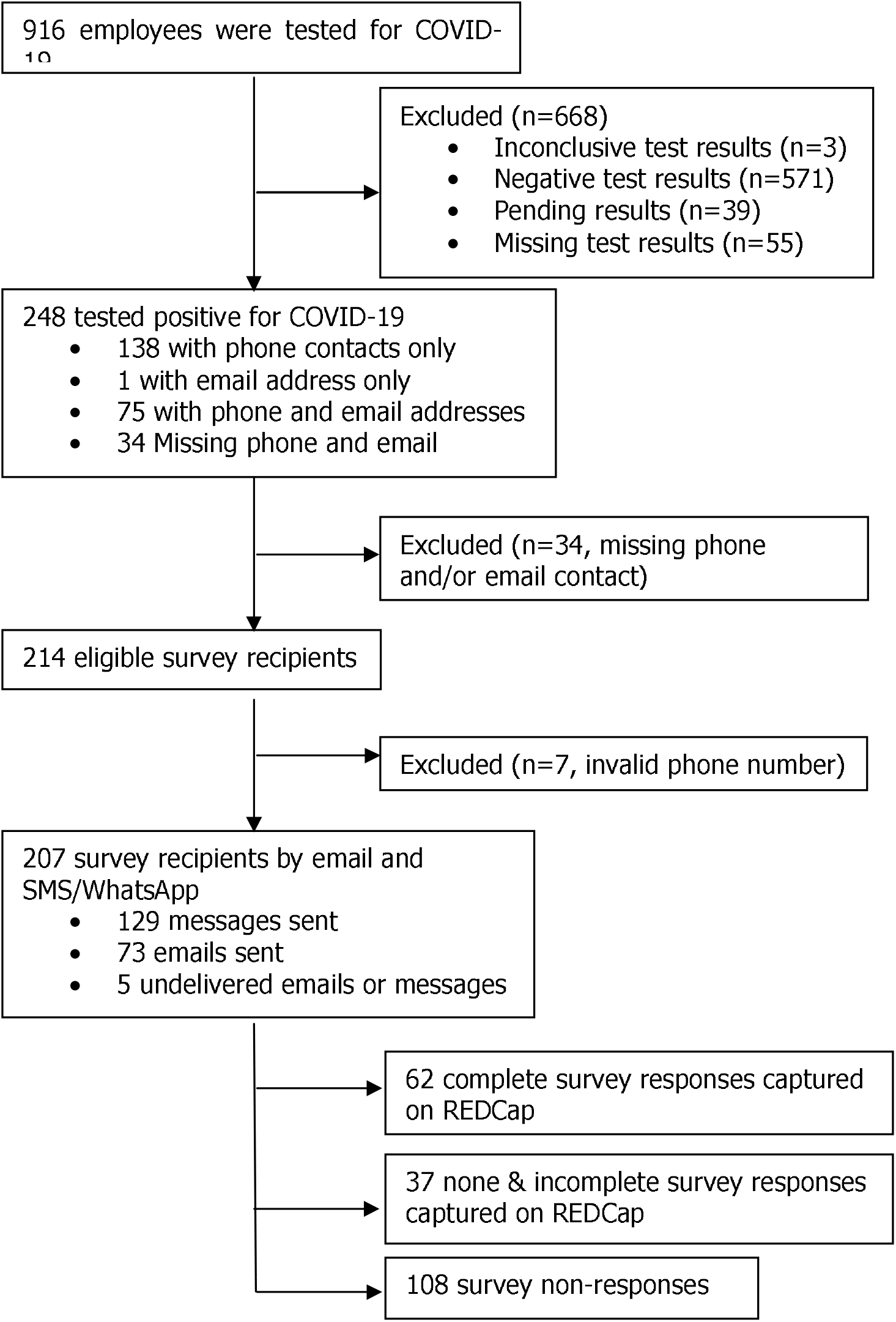
Selection criteria for eligible participants.

**Figure 2:**
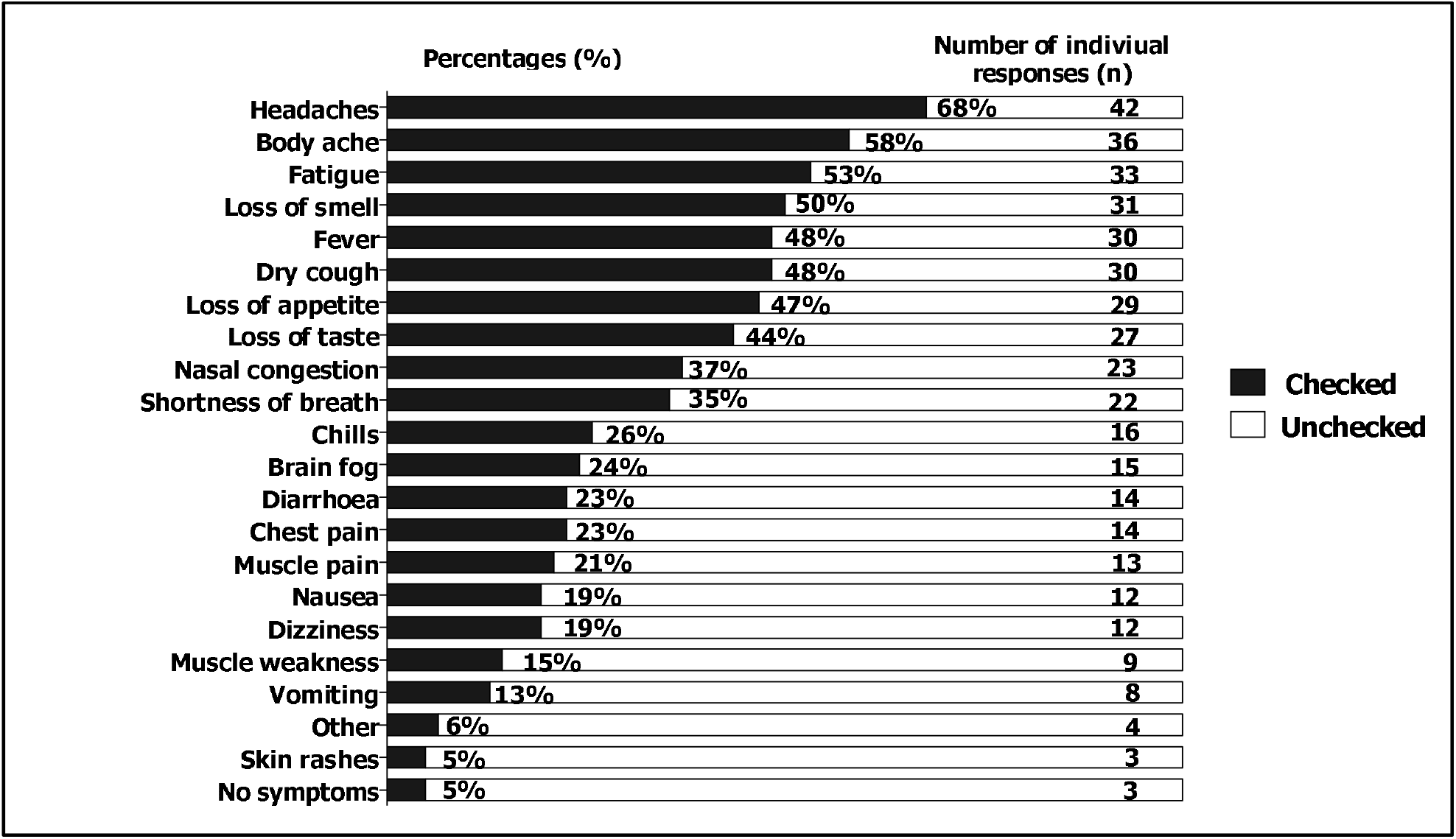
Acute COVID-19 symptoms at time of diagnosis

**Figure 3:**
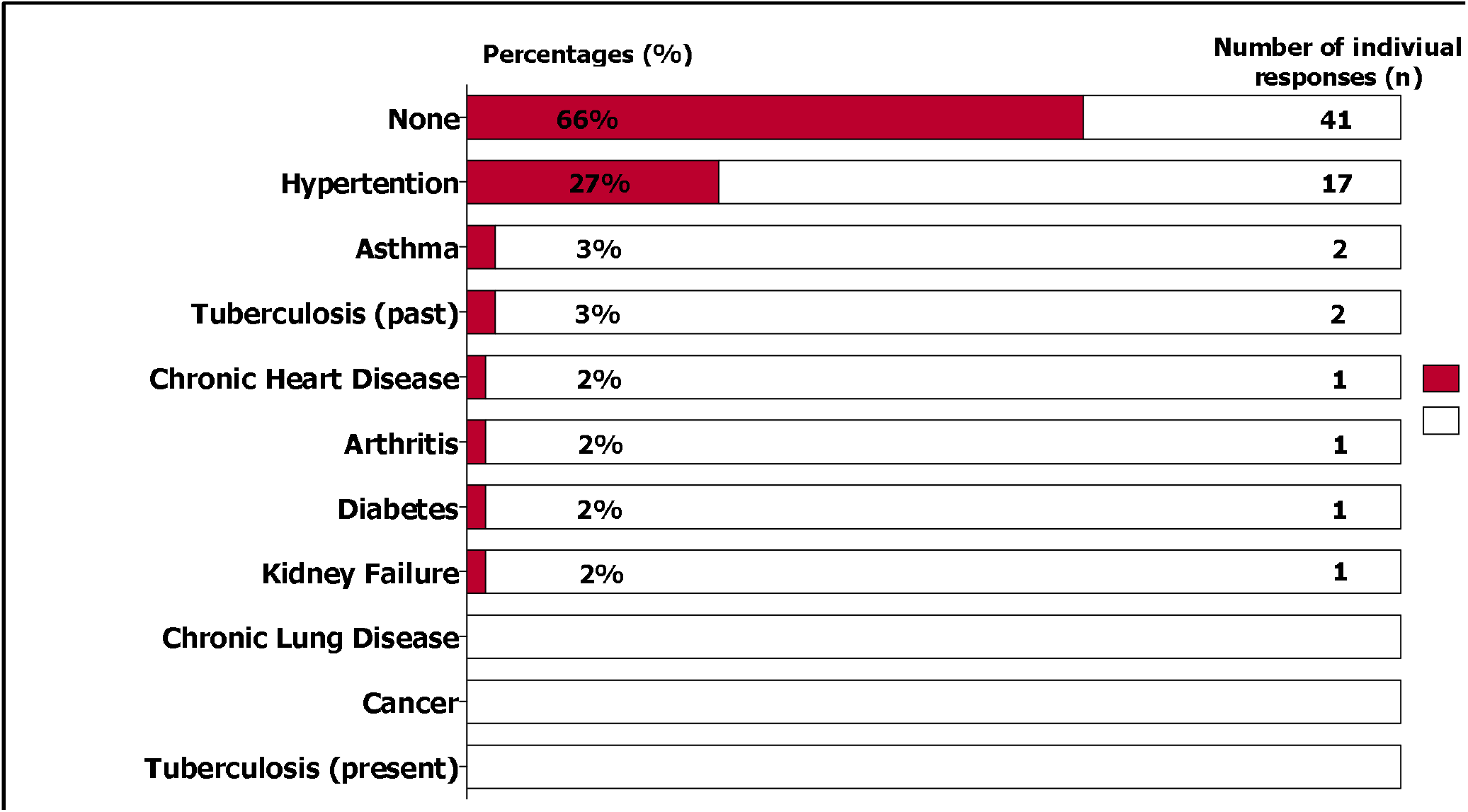
Reported comorbidities among respondents.

The median age of the respondents (n=62) was 33.5 years (Interquartile range {IQR}= 30–44 years), the majority, 31(50%), were between the ages 23–33 years, and 47 (∼76%) were females (Table 1). African (87%; n=54) was the most common ethnicity, followed by coloured (∼10%; n=6). The majority (∼86%; n=53) had never smoked and 6.4% (n=4) were current smokers. Of the four current smokers, each had been smoking <10 cigarettes per day for the past 3, 5, 10 and 14 years, respectively.

**Table 1:** Characteristics of survey participants, N=99

| Demographics | N | Survey Participants |  |
| --- | --- | --- | --- |
|  |  | Respondents<br>n (%) | Non-respondents<sup>‡</sup><br>n (%) |
| **Age (median {IQR})** | 36 (30-34) | 33.5 (30-44) | 37 (31-44) |
| **Sex** |  |  |  |
| Female | 65 | 47 (75.8) | 16 (43.2) |
| Male | 31 | 15 (24.2) | 18 (48.7) |
| Missing | 3 | 0 (0.0) | 3 (8.1) |
| **Age Groups** |  |  |  |
| 23–33 | 45 | 31 (50.0) | 14 (38.0) |
| 34–44 | 30 | 17 (27.4) | 13 (35.0) |
| 45–55 | 15 | 9 (14.5) | 6 (16.2) |
| ≥56 | 6 | 5 (8.1) | 1 (2.7) |
| Missing | 3 | 0 (0.0) | 3 (8.1) |
| **Ethnicity** |  |  |  |
| African | 80 | 54 (87.1) | 26 (70.3) |
| Caucasian | 7 | 2 (3.2) | 5 (13.5) |
| Coloured | 10 | 6 (9.7) | 4 (10.8) |
| Missing | 2 | 0 (0.0) | 2 (5.4) |
| **Smoking Status** |  |  |  |
| Current smoker | 9 | 4 (6.4) | 5 (14.0) |
| Never smoker | 66 | 53 (85.5) | 13 (35.0) |
| Past smoker | 8 | 5 (8.1) | 3 (8.0) |
| Missing | 16 | 0 (0.0) | 16 (43.0) |
| **Alcohol Intake** |  |  |  |
| Daily | 2 | 2 (3.2) | 0 (0.0) |
| Occasional | 32 | 24 (38.7) | 8 (22.0) |
| Not at all | 49 | 36 (58.1) | 13 (35.0) |
| Missing | 16 | 0 (0.0) | 16 (43.0) |
| **Recreational Drug Use** |  |  |  |
| Daily | 2 | 1 (1.6) | 1 (2.7) |
| Occasional | 1 | 1 (1.6) | 0 (0.0) |
| Not at all | 80 | 60 (96.8) | 20 (54.1) |
| Missing | 16 | 0 (0.0) | 16 (43.2) |
| **Total** | **99** | **62** | **37** |

<sup>‡</sup> includes partial and incomplete respondents captured on REDCap

### Clinical manifestations and management of acute COVID-19 infection

Symptoms reported at time of diagnosis included but were not limited to headache (68%; n=42), body ache (58%; n=36), fatigue (53%; n=33), loss of smell (50%; n=31), dry cough (48%; n=30), fever (48%; n=30), loss of appetite (47%; n=29), and loss of taste (44%; n=27) (Fig.2). While 66% (n=41) reported no underlying medical condition, hypertension (27%; n=17) was most common among those who reported a comorbid condition (Fig.3), particularly in females (n=13; 76.55%) and those aged 45–55 years (41%; n=7).

In terms of COVID-19 management after diagnosis, 55 (88.7%) self-isolated, 7 (11.3%) were admitted in hospital, 1 (1.6%) was admitted in a general ward for six days, 3 (4.8%) were in intensive care unit (ICU) for three, five and six days each. For hospital admissions, none was on ventilator, diagnosed with stroke nor received dialysis for kidney failure, and one (1.6%) was sedated.

### Clinical symptoms post-acute COVID

Following post-acute COVID-19 infection, fatigue (42%), anxiety (34%), difficulty sleeping (31%), chest pain (24%), muscle pain and brain fog (21% each) were the six major symptoms experienced by staff. Joint pain was experienced by 19% of respondents (Fig. 4). The duration of symptoms varied between one week to more than three months. Except for shortness of breath, all other symptoms were experienced for longer than a week (Supplementary Fig.1).

**Figure 4:**
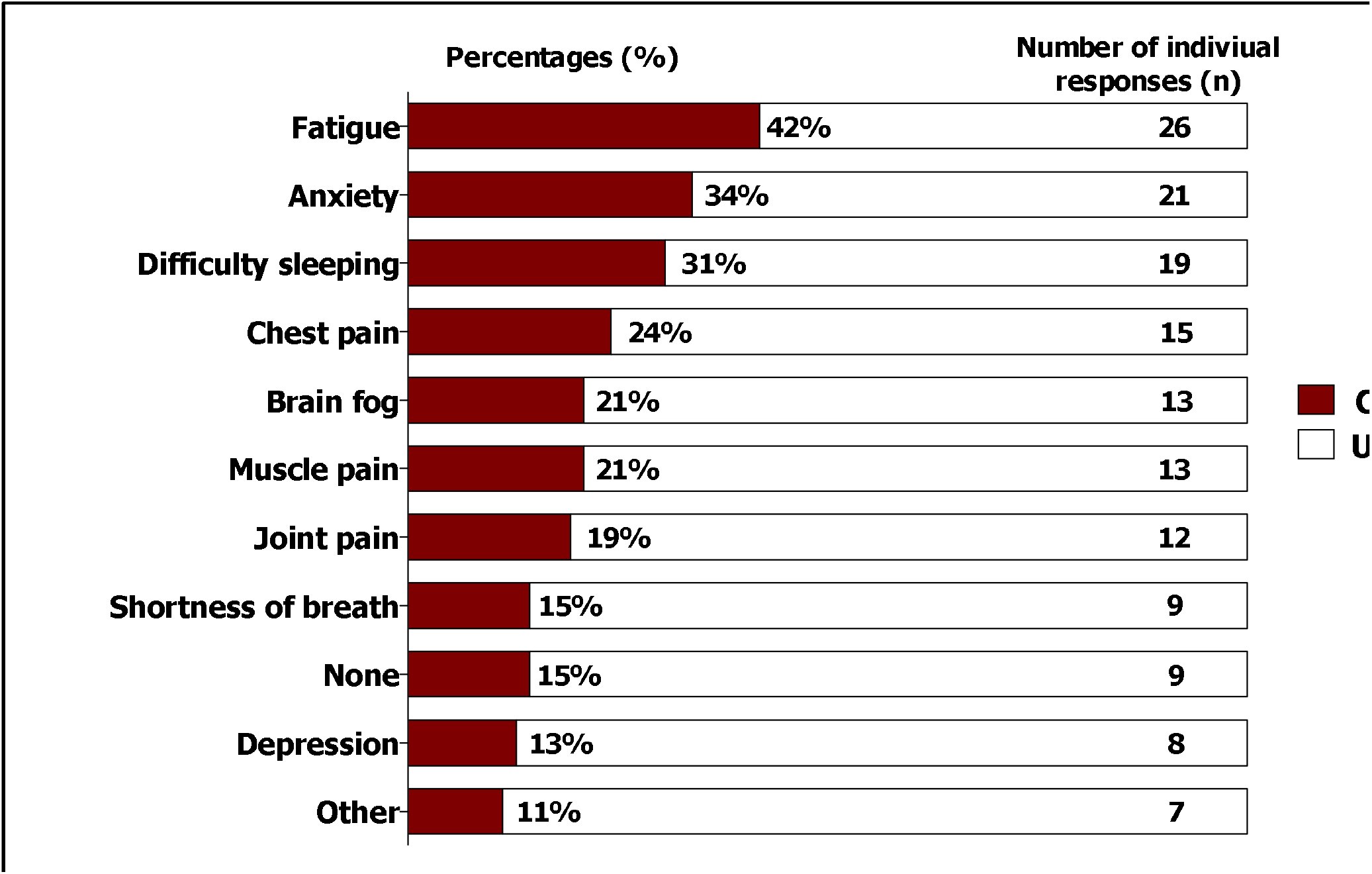
Reported post-acute COVID-19 symptoms among respondents.

At time of survey, fatigue (18%, n=11), anxiety (15%; n=9), difficulty sleeping (13%; n=8), were the three most frequently reported symptoms (Fig. 5) and three most bothersome symptoms (Supplementary Fig.2)

**Figure 5:**
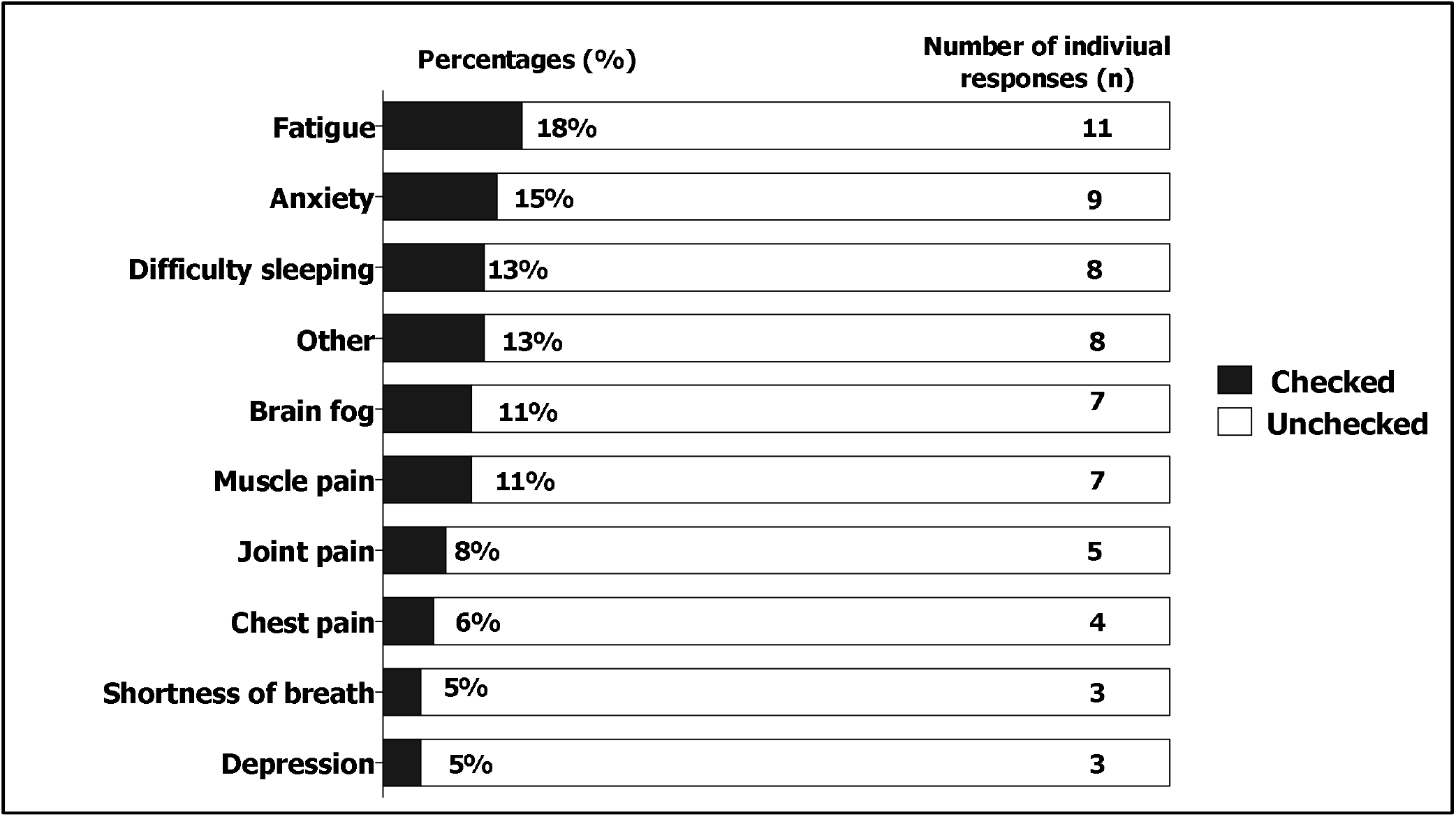
Reported post-acute COVID-19 symptoms at time of survey among respondents.

The number of respondents who participated in some of form of moderate to strenuous physical activity were 45 (73%) before COVID-19 diagnosis and 38 (61%) post-acute COVID-19. Physical activity was performed at a mean of 3.4 (SD=1.7) days (95% CI: 2.6 – 4.0) before and 2.9 (SD=1.5) days (95% CI: 2.3 – 3.4) after COVID-19 recovery. Similarly, physical activity was performed at a mean of 32.9 (SD=22.8) minutes (95% CI: 25.0 – 40.9) before and 29.4 (SD=19.0) minutes (95% CI: 22.8 – 36.0) post-acute COVID-19.

Persistent (Long) COVID-19 (symptoms experienced for four weeks and longer) was observed in 35.5% (22 out of 62) of the respondents. Of these, the median age was 38.5 years (IQR= 36– 46 years as opposed to 31.5 years (IQR= 29–41 years) for those without Long COVID, 36% (8) were aged 34–44 years, and 68% (15) were female (Table 2). While there was an observed significant difference in age between staff with Long COVID and those without, there were no statistical differences by sex, ethnicity, smoking, alcohol and drug intake. Physical activity was not significantly impacted by Long COVID.

**Table 2:** Characteristics of survey respondents with Long COVID-19

| Demographics | N | Long COVID-19 |  | p-Value |
| --- | --- | --- | --- | --- |
|  |  | No<br>n (%), 40 (65.5) | Yes<br>n (%), 22 (34.5) |  |
| **Median Age (IQR)** | 33.5 (30–44) | 31.5 (29–41) | 38.5 (33–46) |  |
| **Age Groups** |  |  |  | **0.045\*** |
| 23–33 | 31 (50.0) | 25 (62.5) | 6 (27.3) |  |
| 34–44 | 17 (27.4) | 9 (22.5) | 8 (36.4) |  |
| 45–55 | 9 (14.5) | 4 (10.0) | 5 (22.7) |  |
| ≥56 | 5 (8.1) | 2 (5.0) | 3 (13.6) |  |
| **Sex** |  |  |  | 0.298 |
| Female | 47 (75.8) | 32 (80.0) | 15 (68.2) |  |
| Male | 15 (24.2) | 8 (20.0) | 7 (31.8) |  |
| **Ethnicity** |  |  |  | 0.087* |
| African | 54 (87.1) | 37 (92.5) | 17 (77.3) |  |
| Caucasian | 2 (3.2) | 0 (0.0) | 2 (9.1) |  |
| Coloured | 6 (9.7) | 3 (7.5) | 3 (13.6) |  |
| **Smoking Status** |  |  |  | 0.141* |
| Current smoker | 4 (6.4) | 4 (10.0) | 0 (0.0) |  |
| Never smoker | 53 (85.5) | 34 (85.0) | 19 (86.4) |  |
| Past smoker | 5 (8.1) | 2 (5.0) | 3 (13.6) |  |
| **Alcohol Intake** |  |  |  | 0.099* |
| Daily | 2 (3.2) | 0 (0.0) | 2 (9.1) |  |
| Occasional | 24 (38.7) | 18 (45.0) | 6 (27.3) |  |
| Not at all | 36 (58.1) | 22 (50.0) | 14 (63.6) |  |
| **Recreational Drug Use** |  |  |  | 0.122* |
| Daily | 1 (1.6) | 0 (0.0) | 1 (4.5) |  |
| Occasional | 1 (1.6) | 0 (0.0) | 1 (4.5) |  |
| Not at all | 60 (96.8) | 40 (100.0) | 20 (91.0) |  |
| **Total** | **62** | **40** | **22** | **62** |

\*p-value obtained with Fischer's exact test.

## Discussion

In this survey, we aimed at identifying the persistent physical and/or mental health symptoms that Right to Care (RTC) employees who have past the acute phase of COVID-19 continue to experience. Our findings showed that more than a third of the respondents had persistent symptoms for four weeks and beyond. The persistent symptoms post-acute COVID-19 is not uncommon. Similar findings have been reported elsewhere.^[9,10]^ Compared to other studies,^[11,12]^ our sample population consisted mostly of non-hospitalized individuals.

We found that all frontline cadres had been positively affected by COVID-19 since its appearance in South Africa. However, those highly impacted had been community health workers and medical/clinical personnel. This is concurrent with national COVID-19 statistics as well as elsewhere.^[13]^

In our survey, hypertension was a common comorbid condition among respondents and 47% reported persistence in at least one symptom, particularly fatigue, anxiety, difficulty sleeping, muscle pain and brain fog. Consistent with existing findings, fatigue was the most reported persistent and bothersome symptom post-acute SARS-CoV-2 infection.^[14,15]^ Although hypertension has been reported as a significant predictor for mortality in patients with COVID-19, ^[4]^ our study revealed no statistically significant association of hypertension with fatigue or the other reported persistent symptoms.

We also found that there was no association of sex with fatigue as opposed to the findings from a post-acute COVID-19 Chinese study, which suggested sex differences.^[4]^ Women were more likely to experience fatigue and anxiety/depression at 6□months of follow-up. Regarding level of physical activity, we found that for a minority of respondents the level of physical activity reduced in frequency and duration. This could be as a result of the lockdown regulations put in place in the first and second wave of the infection^[16,17]^ not necessarily because of the presence of persistent symptoms.

There are a couple of limitations to this survey. Firstly, the small sample of positives for SARS-CoV-2 diagnostic test result does not provide enough statistical power to determine risk factors for Long COVID. However, the survey was able to answer the main survey question. Our study has shown the impact on physical and mental health that COVID-19 has among infected individuals even in the absence of active SARS-CoV-2 virus. Our population was mostly health workers who are in the frontline dealing with the pandemic above other existing infections such as TB and HIV. As such, the impact of COVID even among those that are beyond the active phase of infection could have direct impact in health service delivery. Given the rise in cases of COVID-19 in South Africa and the world at large, the prevalence of Long COVID is likely to be substantial and therefore need for rehabilitation programs targeted at each of the persistent (Long) COVID symptoms is critical. It highlights the importance of slowing the spread of COVID-19 through validated public health measures and vaccinations and continued research into Long COVID to find rehabilitation/treatment options. While our study numbers were small, future work should include detailed physical and mental examination to guide patient management.

## Data Availability

All data produced in the present study are available upon reasonable request to the authors

## Acknowledgments

We would like to thank all respondents for their time and energy in contributing to the study and helping to refine the survey. We would like to thank Andres Montaner for his assistance in providing the data, and Sheila Stanford for assisting with outreach on email. We would like to also thank Tafadzwa Muzvidziwa and team for their help with IT support.

## Author contributions

CSC, SH, ALS, FL, and FA conceptualized the project. CSC, SH, ALS and CWK designed the survey. CWK cleaned, analysed the quantitative data, performed the statistical analyses and wrote the manuscript. All authors reviewed and approved the manuscript for publication. The corresponding author attests that all listed authors meet authorship criteria and that no others meeting the criteria have been omitted.

## Competing Interest

None declared

**Supplementary Table 1:** Characteristics of RTC employees who tested for COVID-19

| Variables | N | COVID-19 Test Outcome |  | p-value |
| --- | --- | --- | --- | --- |
|  |  | Positive n (%) | Other n (%) |  |
| **Staff Cadres** |  |  |  | **0.001** |
| Community/Field health Workers | 329 | 95 (38.3) | 234 (35.0) |  |
| Medical/Clinical personnel | 137 | 42 (16.9) | 95 (14.2) |  |
| Data capturers/Clerks | 140 | 32 (13.0) | 108 (16.2) |  |
| Programme Coordinators | 25 | 5 (2.0) | 20 (3.0) |  |
| Programme Support Staff | 82 | 35 (14.1) | 47 (7.0) |  |
| Missing | 203 | 39 (15.7) | 164 (24.6) |  |
| **Reasons for testing** |  |  |  | **0.010\*** |
| COVID-19 contact | 271 | 73 (29.4) | 198 (29.6) |  |
| COVID-19 PUI/Suspect | 503 | 151 (60.9) | 352 (52.7) |  |
| Routine Health Surveillance | 125 | 24 (9.7) | 101 (15.1) |  |
| Travel requirements | 4 | 0 (0.0) | 4 (0.6) |  |
| Missing | 13 | 0 (0.0) | 13 (2.0) |  |
| **Province** |  |  |  | **0.001\*** |
| Gauteng | 62 | 16 (6.5) | 46 (6.9) |  |
| Free State | 326 | 81 (33.0) | 245 (36.7) |  |
| Mpumalanga | 249 | 61 (25.0) | 188 (28.1) |  |
| Eastern Cape | 24 | 15 (6.1) | 9 (1.4) |  |
| Northern Cape | 2 | 2 (1.0) | 0 (0.0) |  |
| Western Cape | 248 | 70 (28.2) | 178 (26.7) |  |
| North West | 5 | 3 (1.2) | 2 (0.3) |  |
| **Districts** |  |  |  | **0.000\*** |
| City of Johannesburg MM | 33 | 8 (3.2) | 25 (4.0) |  |
| City of Tshwane MM | 23 | 7 (2.8) | 16 (2.4) |  |
| Lejweleputswa | 21 | 6 (2.4) | 15 (2.3) |  |
| Ehlanzeni | 240 | 58 (23.4) | 182 (27.3) |  |
| Gert Sibande | 11 | 2 (0.8) | 9 (1.4) |  |
| Nelson Mandela Bay MM | 5 | 3 (1.2) | 2 (0.3) |  |
| Amatole | 11 | 7 (3.0) | 4 (1.0) |  |
| OR Tambo | 5 | 5 (2.0) | 0 (0.0) |  |
| Thabo Mofutsanyane | 302 | 73 (29.4) | 229 (34.3) |  |
| Dr Kenneth Kaunda | 7 | 6 (2.4) | 1 (0.2) |  |
| Central Karoo | 104 | 15 (6.1) | 39 (6.0) |  |
| Cape Winelands | 54 | 15 (6.1) | 39 (6.0) |  |
| Overberg | 86 | 25 (10.0) | 61 (9.1) |  |
| City of Cape Town MM | 2 | 0 (0.0) | 2 (0.3) |  |
| Ngaka Modiri Molema | 1 | 1 (0.4) | 0 (0.0) |  |
| Nkangala | 2 | 1 (0.4) | 1 (0.2) |  |
| Missing | 9 | 1 (0.4) | 8 (1.2) |  |
| **Total** | **916** | **248** | **668** |  |

\*p-value obtained with Fischer's exact test.

**Supplementary Figure 1:**
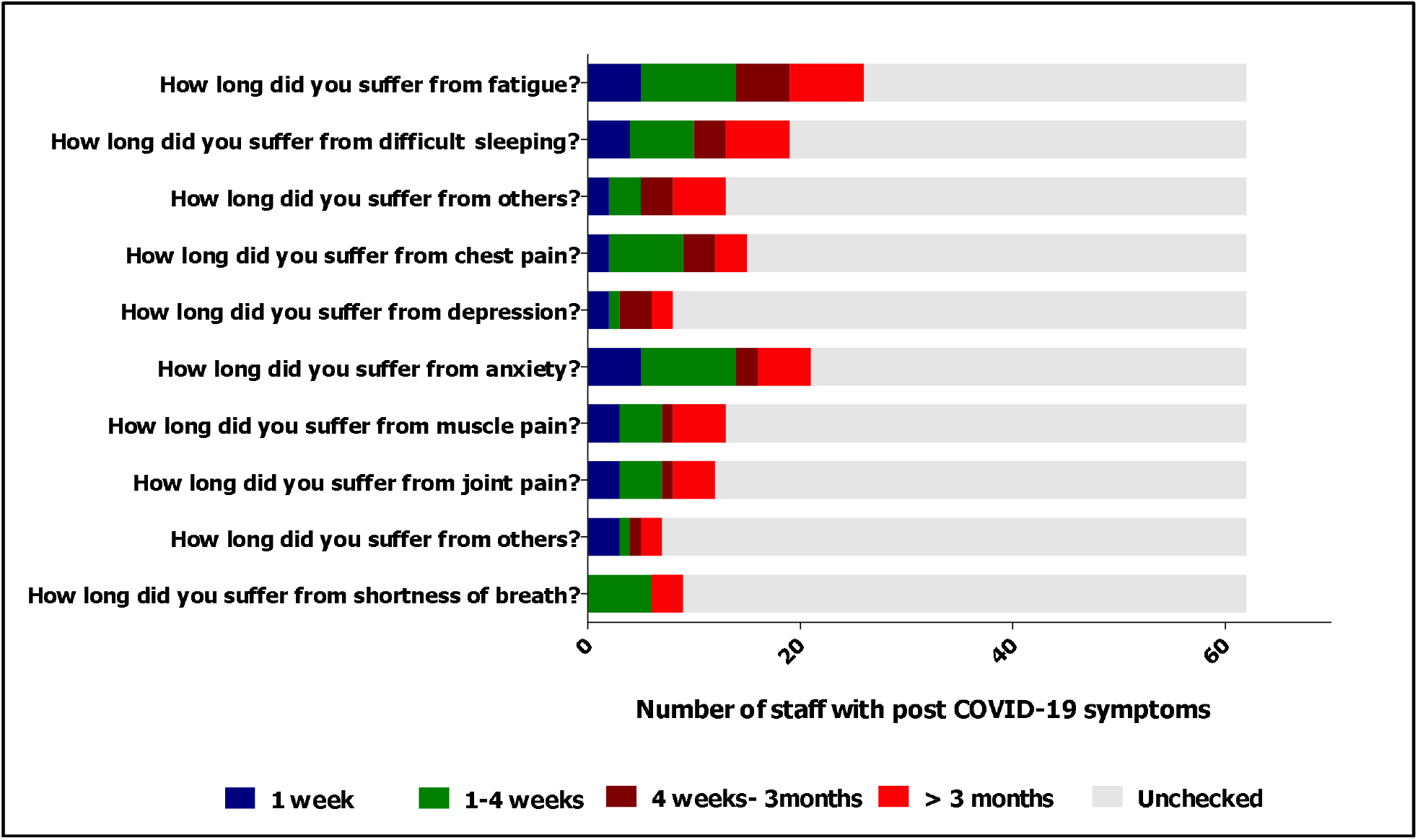
Duration of post-acute COVID-19 symptoms among survey respondents

**Supplementary Figure 2:**
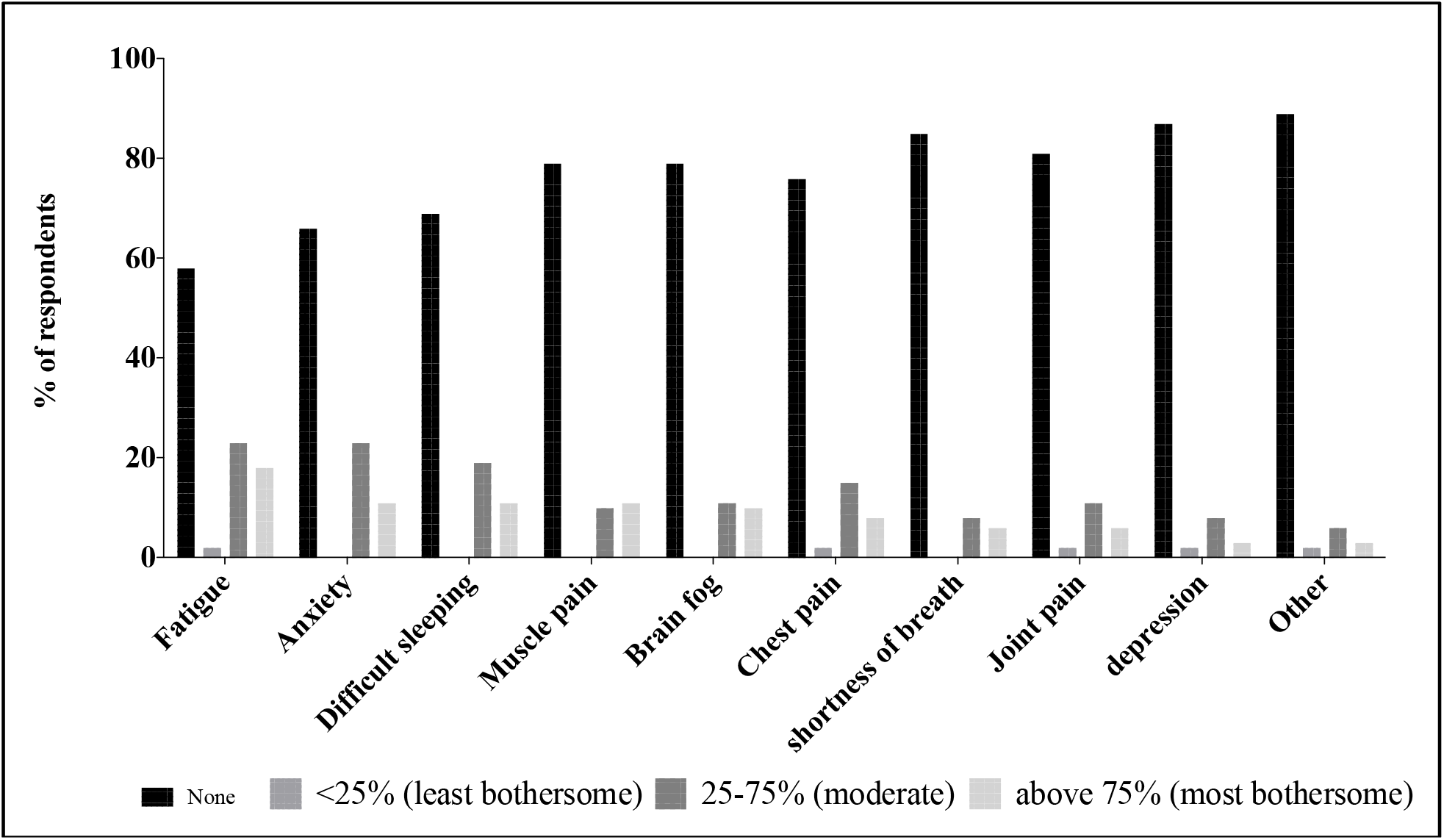
Reported bothersome/troubling post-acute COVID-19 symptoms among respondents.

